# What can we learn from talented General Practitioners and Elderly Care Medicine Specialists? A protocol for a qualitative interview study

**DOI:** 10.1101/2022.11.28.22282747

**Authors:** Abdullah Khawar, Mechteld Visser, Marianne Mak, Martin Smalbrugge, Irene Slootweg, Nynke van Dijk

## Abstract

1.

**Background:** Some postgraduate medical trainees in the General Practice (GP) and Elderly Care Medicine (ECM) training program, are considered being talented trainees (TTs) by their peers, teachers (at the institute), trainers (in clinical practice), and/or patients. We are currently unaware if these TTs are trained to meet their maximal potential or whether they are not fully stimulated during their learning process in the postgraduate training program. It is important to acknowledge them, because if we fail to acknowledge and suitably challenge TTs, it may lower their work ethos and satisfaction or cause loss of our TTs. As a first step, we will explore the way TTs make use of the learning possibilities provided during the postgraduate training program and what stimulates and hinders their learning in the clinical workplace.

Knowledge on this subject is important for two reasons. First, knowing how TTs learn, can provide insights for improving learning in the workplace for all trainees. Secondly, with this knowledge, we can enrich the development of TTs. Thirdly, enhancement of workplace-based learning might lead to higher quality of learning, resulting in health care professionals trained to their full potential.

**Methods and analysis:** We will address these questions in an open approach, conducting explorative qualitative semi-structured interview study with GPs and ECM specialists, who during their postgraduate training were seen as talented by their third year supervisors and completed their postgraduate medical training two or less than two years ago. We will perform the interviews in Dutch. We expect that the interview will not take longer than 45-60 minutes per interview. We plan to start at the beginning of 2023 and will continue until data sufficiency is reached. All interviews will be audio-recorded and transcribed verbatim. We will use thematic analysis to analyse the transcripts of the interviews. This is an iterative process of familiarizing yourself with the data, generating initial codes, searching for themes, reviewing themes, defining and naming themes. We will use MAXQDA 2022 software.

**Ethical considerations:** The Ethics committee of the Dutch Association of Medical Education (NVMO) gave ethical approval for this work (NERB dossier number: 2022.7.3).

**Data availability statement:** All data produced in the present study are available upon reasonable request to the authors.

**Funding statement:** This study is funded by ZonMw (project number 839130008).

## 2. Introduction

From experience, some postgraduate medical trainees in the General Practice (GP) and Elderly Care Medicine (ECM) training program, are considered being talented or excellent trainees by their peers, teachers (at the institute), trainers (in clinical practice), and/or patients. However, we are currently not equipped to acknowledge and suitably challenge talented or excellent postgraduate medical trainees in the postgraduate medical GP and ECM training program.

The learning of postgraduate medical trainees has been subject of much research. Postgraduate medical trainees learn in the clinical workplace, which is also known as workplace-based learning. The clinical workplace is a rich environment to learn from experience (1, 2) and from others (1). Learning occurs through engaging in work-related activities, by which postgraduate medical trainees interpret and construct meaning and eventually expand their knowledge (3). They do so under supervision of their clinical trainer. There are many learning theories that address aspects of workplace-based learning, for example self-regulated learning (4, 5) deliberate practice(6), adaptive expertise(7) and socio-cultural theory of learning(8, 9).

Our knowledge about how good clinical students learning in the workplace has grown. In the undergraduate medical setting, good clinical students are described as being ‘proactive in seeking out relevant learning experiences, enthusiastic, and motivated’(10) and ‘seeking opportunities for improvement and growth’(11). Furthermore, ‘experienced’ undergraduate medical students, who learn in a clinical setting, seem to be better situated in the clinical environment, compared to their novice peers. They employ self-regulated learning, get a grip on their learning, understand their role and have an idea what they want to become(12). In the postgraduate setting, it is unclear how the specific group of talented postgraduate medical trainees learn, how they regulate or manage their learning in clinical practice and develop during their postgraduate training program.

Also, we are currently unaware if these talented trainees are trained to meet their maximal potential or whether they are under stimulated during their learning process in the postgraduate training program. It may well be that we train all trainees in a ‘one-size-fits-all’ with our current postgraduate training program. This is troublesome, as we know that it is important to acknowledge them, because if we fail to acknowledge and suitably challenge talent, it may cause loss of our talented trainees and lowers their work ethos(13). As a first step in investigating if and how we best can stimulate talented trainees to their best potential, we will explore the way talented trainees learn and what stimulates and hinders their learning in the clinical workplace.

We will address these questions in an open approach, conducting explorative qualitative semi-structured interview study with GPs and ECM specialists, who during their postgraduate training were seen as talented by their supervisors.

## 3. Research aims

In order to acknowledge and suitably challenge talented or excellent postgraduate medical trainees in the postgraduate medical GP and ECM training program, we aim to answer the following research questions:

1. How did talented trainees learn during their postgraduate medical training program?
2. What did help and hinder talented trainees during their development?

## 4. Target audience

Our target audience will consist of GPs and the ECM specialists, who were considered being talented during their postgraduate training program by their supervisors of the last training year. For detailed information on recruitment, see *7*.*1 recruitment*.

In the Netherlands, eight academic centres provide the postgraduate medical training for GPs, these include Academic Medical Centre (AMC), Vu Medical Centre (VuMC), University Medical Centre Groningen (UMCG), Maastricht University Medical Centre+ (MUMC+), Radboud University Medical Centre (Radboudumc), Erasmus Medical Centre (EMC) and Leiden University Medical Centre (LUMC), UMC Utrecht (UMCU), and three centres provide the postgraduate medical training program for ECM specialists, which include the Vu Medical Centre (VuMC), Leiden University Medical Centre (LUMC), and Radboud University Medical Centre (Radboudumc).The GP and ECM postgraduate medical training programs have similar training settings, in which the first and third year take place in a general practice or elderly care facility, respectively, and a second year that consists of hospital internships for both postgraduate training programs. During the first and third year, training mostly takes place in the clinical workplace (4 days/week in the practice, 1 day/week of formal education).

We want to perform this study with GP and ECM specialists, because, we believe that asking current postgraduate medical trainees to participate in our study is unethical as it can lead to the imposition of unintended differences between the current postgraduate medical trainees, in which one postgraduate medical trainee is earmarked talented and the other is not.

## 5. Design and Procedure

### 4.1 Research design

We will conduct an explorative qualitative semi-structured interview study with GPs and ECM specialists (14). We will adopt a constructivist paradigm and use an interview protocol with topics to guide the interviews. This interview protocol will consist of open questions for the interviewees, allowing respondents to describe in their own words the processes or opinions to specific subjects as formulated by the researcher, and giving room for emerging questions during the interview(15).

### 4.2 Research procedure

First, an interview protocol will be produced using literature on learning of postgraduate medical trainees.

#### Part 1. Testing interview procedure

We will perform a test to ensure that the questions, information provided and the interview procedure is clear, and to improve our interview protocol and procedure. For this test, we will recruit 3-5 participants including both medical trainees from the GP training as ECM medical trainees. Also, someone with knowledge of performing an interview study will be included.. We will send all information to our test participants. Then, will perform the interview, as we would with the potential participants of this study. Afterwards, we will ask the interviewee for feedback on the procedure, information provided, and interview. Because of the restricted aim of this pilot study interviews will not be recorded. The results will help us to optimize the interview topics and procedure.

#### Part 2. Interview study with GP and ECM specialists

The interviews will take place, after receiving written informed consent, at a location and time favourable to the interviewees. This could be either face-to-face or online. In case of an online interview, we will make use of Microsoft Teams. This is because we have a secure account, to ensure the privacy of the participant. We will perform the interviews in Dutch. We expect that the interview will not take longer than 45-60 minutes per interview. We plan to start at the beginning of 2023 and will continue until data sufficiency is reached(16).

The provisional topic list of our interview will consist of questions concerning learning, and factors related to what helps and hinders (see *attachment 8*.*1*).

## 6. Methods

### 5.1 Outcome(s)

Our primary outcomes are insights into the learning strategies of talented GPs and ECM specialists, and what helped and hindered talent trainees in their development.

### 5.2 Interim termination of participation

If participants want to withdraw or quit after commitment to participate, even if the interviews have taken place, we will remove their data, and, if the analysis has not yet been started, we will not use their data in this study, as indicated in the information letter. After analysis it is impossible to relate codes and themes to individual statements, and therefore we cannot remove the data after analysis has started.

## 7. (Statistical) analyses

We will collect the following demographic data: age, gender, years since completion of postgraduate medical training and if they are a GP or ECM specialist.

All interviews will be audio-recorded and transcribed verbatim. We will use thematic analysis to analyse the transcripts of the interviews (17, 18). This is an iterative process of familiarizing yourself with the data, generating initial codes, searching for themes, reviewing themes, defining and naming themes. We will use MAXQDA 2022 software(19).

The initial analysis will be performed by two researchers, who will, independently, produce a code tree based on the first transcript. We will discuss, change and refine the code tree, in an iterative fashion, until we have reached consensus about the final code tree. If disagreement occurs, we will include a third independent research from our research team. Based on the discussion sessions, codes and themes can be refined, changes, added, and/or removed. After consensus, one researchers will continue independently to analyse the other transcripts, using the code tree.

Throughout the analysis and research team discussions, we will keep an audit trail, to provide a clear account of procedures used and choices made.

We will not use any theoretical framework beforehand to analyse or code our data, as we are do not want to take a position in advance on which learning strategies are employed by talented trainees.

We do not expect any missing data, as in this qualitative research approach no specific answers from the interviewees are requested.

## 8. Ethical aspects

### 7.1 recruitment

We aim to include twenty GPs and ECM specialists from each institute. To participate, we use the following criteria: 1) the institute view them as talented/excellent/high-performing, based on the judgement of trainers and teachers, 2) they have completed their postgraduate medical training five or less than five years ago. We believe that if a GP or ECM specialist has completed the postgraduate training program more than a period of five years, it is possible that the postgraduate medical training program has changed to such an extent that the input will not be sufficient for this research.

We intend to approach all institutes, with a GP and/or a ECM postgraduate training program, to recruit potential participants for this interview study. We will ask the heads of the GP and ECM postgraduate training program for permission to include potential participants. We will do so by reaching out to Huisartsopleiding Nederland (HON), via Judy van Es (head of the GP postgraduate training program at Amsterdam UMC, location AMC) and Specialist Ouderengeneeskunde Opleiding Nederland (SOON), via Martin Smalbrugge (head of the ECM postgraduate training program at Amsterdam UMC, location VuMC).

For this research we are interested in current GPs and ECM specialists, who have already completed their training program, and were at the time of their training considered talented by their third-year trainer. In order to reach the group of talented GPs and ECM specialists, as we do not directly have any contact information of this group, we will approach trainers from the third-year of the postgraduate GP and ECM training program from institutes mentioned under heading *3. Target audience*. We will approach those trainers via a contact person at the institutes, and ask the trainers whether they have supervised a talented trainee during the third-year. Talent is difficult to describe precisely and could be subject to subjectivity depending on the assessor. However, to help the trainers deciding whom they could consider as talented we will provide them with a general description, concerning competencies and motivation, and is based on our findings from our most recent systematic review (accepted for publication). The description is as follows; ‘the GP/ECM specialist is talented because he/she was talented in several competencies (namely, medical knowledge, communication and professionalism), and is clearly motivated for patient care and to learn’.

With this description we will ask third-year trainers to approach the potential candidates and ask if they want to participate in this interview study. We will ask the trainer to forward a standardized e-mail for participating in our interview study, which includes the participant information letter and informed consent form (see *attachment 8*.*2, attachment 8*.*3* and *attachment 8*.*4*). Potential candidates can either send an e-mail to contact the first author (Abdullah Khawar) via or they give permission to the third-year trainer to share their e-mail address with Abdullah Khawar.

We will contact the participant via telephone or e-mail, depending on the participant’s preference. We will shortly address the goal, research question and procedure, and check if this is clear with the potential participant. If he/she agrees on participation, we will then plan our interview, as stated earlier, on a location favourable to the participant. This can be either in real life or digital.

The participant will then receive a confirmation e-mail of the appointment (*attachment 8*.*5*), and again we will send the participant information letter and informed consent form (*attachment 8*.*3* and *attachment 8*.*4*), if we have not yet received their signed informed consent form. Written informed consent must be obtained before start of the interview. Without a signed informed consent by both parties (i.e. researcher or representative, and participant), no interview will take place.

If we have insufficient participants for this study, we will actively promote this study and ask for participation, by attending physical meetings.

### 7.2 Information and consent

#### 7.2.1 Pros and cons and possible risk of the study

Pros:

- The participants are GPs and ECM specialists, and therefore they do not have any ties to the institute, related to assessment of their performance;

Cons:

- There is an expected maximum time investment of two hours;
  - A maximum of thirty minutes of reading an replying to the first mail and initial contact via the telephone or e-mail
  - A maximum of sixty minutes for the interview
  - A maximum of thirty minutes of reading and responding to the summary (which can include adjusting, nuancing, and/or providing additional suggestions to the summary) provided after the interview by the researcher
- We do not expect any other cons related to the participation

#### 7.2.2 Termination of participation

A Participant can terminate his/her participation at any time. When withdrawing, the participant should let the research team know of his/her withdrawal, by e-mailing Abdullah Khawar. This to ensure any data collected will be removed. As indicated in *5*.*2 Interim termination of participation*; if the analysis has not yet been started, we will not use their data in this study, as indicated in the information letter. After analysis it is impossible to relate codes and themes to individual statements, and therefore we cannot remove all data after the analysis has started. The participant is not required to give a reason to withdraw. Withdrawal does not lead to any adverse consequences.

#### 7.2.3 Justification of the study

Knowledge on this subject is important for two reasons. First, knowing how they learn, can provide insights for improving learning in the workplace for *all* trainees. Secondly, with this knowledge, we can enrich the development of talented postgraduate medical trainees and enhancement in workplace-based learning might lead to higher quality of learning, resulting in health care professionals trained to their full potential.

#### 7.2.4 Rewards

Participants will not receive a reward for their participation.

#### 7.2.5 Data management

The interviews will be conducted by one researcher (Abdullah Khawar).

Other members of the research group will only have access to the encrypted data (Mechteld Visser, Marianne Mak-van der Vossen, and Nynke van Dijk).

Participants will be assigned a participant number. The interviews will be audio-recorded and transcribed verbatim by Abdullah Khawar, however, any traceable information to the participant will be encrypted (i.e., name, workplace, and any other identifying information). Only the encrypted data will be used for analysis. All data (encrypted and not encrypted) will be saved in a designated file, only accessible via a password by Abdullah Khawar, on the AMC secure network. Sharing any encrypted transcript will happen via the AMC secure network. Demographic information (i.e. experience with training and teaching in years, year of training of the trainee and affiliated faculty, gender, age and function of the interviewee), which will have an impact on the results and are therefore important to use in this study, will be saved separately but will not be encrypted.

We will store the informed consent forms in a locked storage, and also make a digital copy of each informed consent form and store it, along with the other data in a secured file on the AMC network.

After completion of this study the data will be kept for ten years (www.vsnu.nl). The paper documents will be kept at in the general archive of the AMC and digital data will be kept on a secured disk of the Department of General Practice in the AMC. After this period, the data will be destroyed according to AMC protocol. This will be done by the Department of General Practice in collaboration with the information and commutations technology (ICT) department of the AMC.

If participants terminate their participation, at any stage of their participation, any data collected will not be included in the analysis and will be removed and deleted from the database directly after withdrawal.

## 9. Attachments

*8*.*1 Interview protocol (in Dutch)*

*8*.*2 Email_uitnodiging (in Dutch)*

*8*.*3 Deelnemersinformatie (in Dutch)*

*8*.*4 Toestemmingsformulier (in Dutch)*

*8*.*5 Email_bevestiging (in Dutch)*

## Data Availability

All data produced in the present study are available upon reasonable request to the authors.

